# REACT-1 round 10 report: Level prevalence of SARS-CoV-2 swab-positivity in England during third national lockdown in March 2021

**DOI:** 10.1101/2021.04.08.21255100

**Authors:** Steven Riley, Oliver Eales, David Haw, Caroline E. Walters, Haowei Wang, Kylie E. C. Ainslie, Christina Atchison, Claudio Fronterre, Peter J. Diggle, Deborah Ashby, Christl A. Donnelly, Graham Cooke, Wendy Barclay, Helen Ward, Ara Darzi, Paul Elliott

## Abstract

**Background:** In England, hospitalisations and deaths due to SARS-CoV-2 have been falling consistently since January 2021 during the third national lockdown of the COVID-19 pandemic. The first significant relaxation of that lockdown occurred on 8 March when schools reopened.

**Methods:** The REal-time Assessment of Community Transmission-1 (REACT-1) study augments routine surveillance data for England by measuring swab-positivity for SARS-CoV-2 in the community. The current round, round 10, collected swabs from 11 to 30 March 2021 and is compared here to round 9, in which swabs were collected from 4 to 23 February 2021.

**Results:** During round 10, we estimated an R number of 1.00 (95% confidence interval 0.81, 1.21). Between rounds 9 and 10 we estimated national prevalence has dropped by ∼60% from 0.49% (0.44%, 0.55%) in February to 0.20% (0.17%, 0.23%) in March. There were substantial falls in weighted regional prevalence: in South East from 0.36% (0.29%, 0.44%) in round 9 to 0.07% (0.04%, 0.12%) in round 10; London from 0.60% (0.48%, 0.76%) to 0.16% (0.10%, 0.26%); East of England from 0.47% (0.36%, 0.60%) to 0.15% (0.10%, 0.24%); East Midlands from 0.59% (0.45%, 0.77%) to 0.19% (0.13%, 0.28%); and North West from 0.69% (0.54%, 0.88%) to 0.31% (0.21%, 0.45%). Areas of apparent higher prevalence remain in parts of the North West, and Yorkshire and The Humber. The highest prevalence in March was found among school-aged children 5 to 12 years at 0.41% (0.27%, 0.62%), compared with the lowest in those aged 65 to 74 and 75 and over at 0.09% (0.05%, 0.16%). The close approximation between prevalence of infections and deaths (suitably lagged) is diverging, suggesting that infections may have resulted in fewer hospitalisations and deaths since the start of widespread vaccination.

**Conclusion:** We report a sharp decline in prevalence of infections between February and March 2021. We did not observe an increase in the prevalence of SARS-CoV-2 following the reopening of schools in England, although the decline of prevalence appears to have stopped. Future rounds of REACT-1 will be able to measure the rate of growth or decline from this current plateau and hence help assess the effectiveness of the vaccination roll-out on transmission of the virus as well as the potential size of any third wave during the ensuing months.

## Introduction

During the spring of 2021, many European populations are suffering from substantial third waves of the COVID-19 pandemic with high pressure on health care systems [1] resulting in stringent social distancing. In England, during the third national lockdown, hospitalisations and deaths have fallen consistently since early January [2]. However, since the first substantial relaxation of lockdown in England with the opening of schools on 8 March 2021 [3], the rate of decline of new cases has slowed considerably [2]. Large-scale testing of school children and their families using lateral flow devices accompanied the opening of schools [4].

The REal-time Assessment of Community Transmission-1 (REACT-1) study augments routine surveillance data for England by measuring swab-positivity for SARS-CoV-2 in a random sample of the community recruited regardless of symptom status [5]. We present here the results from round 10 of REACT-1 from swabs collected from 11 to 30 March 2021. We compare the results of REACT-1 round 10 to round 9, in which swabs were collected from 4 to 23 February 2021.

## Results

In round 10 we found 227 positives from 140,844 swabs giving an unweighted prevalence of 0.16% (0.14%, 0.18%) and a weighted prevalence of 0.20% (0.17%, 0.23%) (Table 1). This represents a ∼60% reduction from round 9 in which unweighted prevalence was 0.42% (0.39%, 0.45%) and weighted prevalence was 0.49% (0.44%, 0.55%).

**Table 1.**
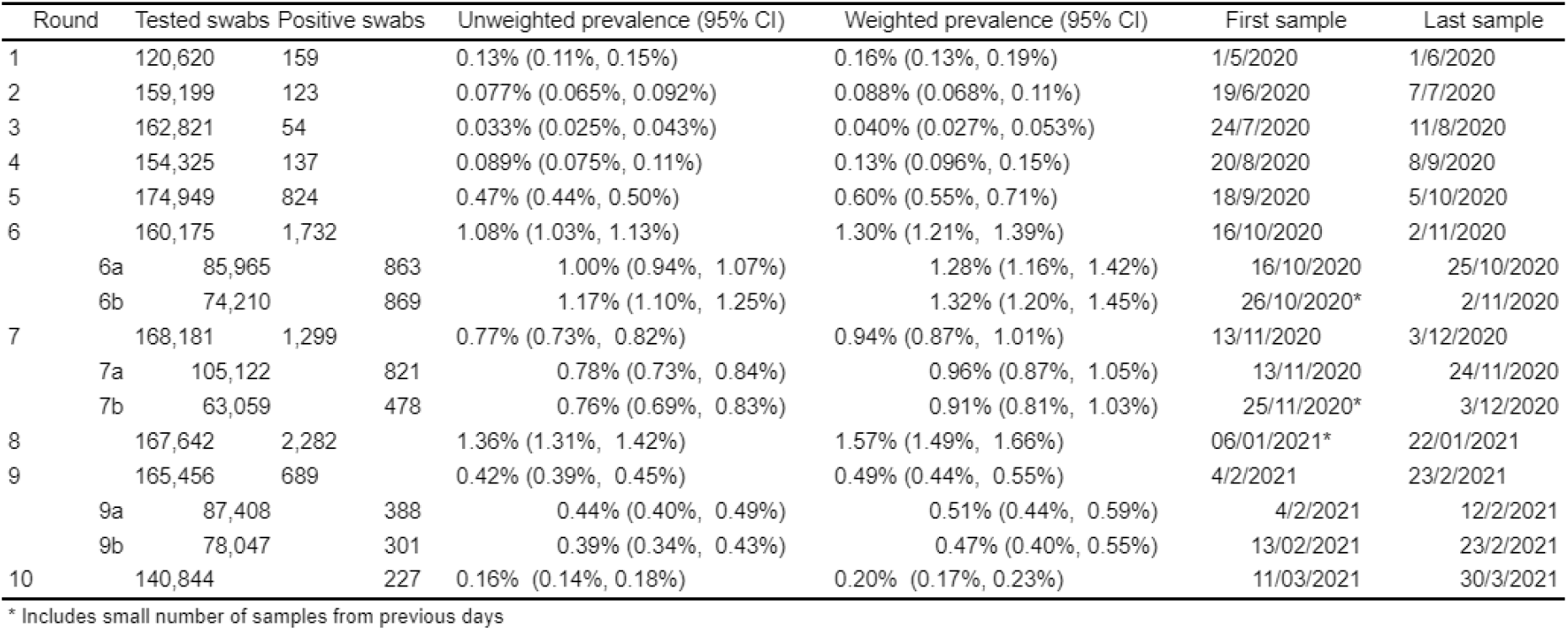
Unweighted and weighted prevalence of swab-positivity across nine rounds of REACT-1 and round 10.

Using a constant growth rate model, on average for England, we found evidence for a decline over the period of round 9 to 10 (Table 2, Figure 1) with an estimated halving time of 26 (23, 31) days and a corresponding R of 0.84 (0.82, 0.86). Within round 10, we estimate an R of 1.00 (0.81, 1.21) with 51% probability that R > 1. These results suggest a slow down in the rate of decline, with prevalence plateauing at around 1 in 500 during round 10 (Figure 1). These results were largely unchanged in sensitivity analyses in which subsets of positive samples were considered: non-symptomatics, only participants with swabs positive for both E and N genes, and only those positive for N gene with a CT value of 35 or below. However, we estimated a lower R of 0.72 (0.50, 0.99) for round 10 based only on the prevalence of positive swabs taken from non-symptomatic participants (Table 2).

**Table 2.** Estimates of national growth rates, doubling times and reproduction numbers for round 9 to round 10, and within round 10.

| Round | Outcome | Growth rate | R | Probability R>1 | Growth(+)/Decay(-) rate |
| --- | --- | --- | --- | --- | --- |
| 9 and 10 | All positives | -0.026 ( -0.031 , -0.022 ) | 0.84 ( 0.82 , 0.86 ) | <0.01 | -26.2 ( -22.6 , -31.0 ) |
|  | Non-symptomatics | -0.020 ( -0.026 , -0.014 ) | 0.88 ( 0.84 , 0.91 ) | <0.01 | -34.8 ( -26.5 , * ) |
|  | Positive for both E and N genes | -0.033 ( -0.039 , -0.027 ) | 0.80 ( 0.77 , 0.83 ) | <0.01 | -20.9 ( -17.7 , -25.3 ) |
|  | Positive for both E and N genes or positive only for N gene with CT 35 or less | -0.026 ( -0.030 , -0.021 ) | 0.84 ( 0.82 , 0.87 ) | <0.01 | -27.0 ( -23.0 , -32.6 ) |
| 10 | All positives | 0.000 ( -0.031 , 0.032 ) | 1.00 ( 0.81 , 1.21 ) | 0.51 | * ( -22.4 , 21.9 ) |
|  | Non-symptomatics | -0.047 ( -0.095 , -0.002 ) | 0.72 ( 0.50 , 0.99 ) | 0.02 | -14.7 ( -7.3 , * ) |
|  | Positive for both E and N genes | -0.009 ( -0.056 , 0.036 ) | 0.94 ( 0.68 , 1.25 ) | 0.35 | * ( -12.5 , 19.1 ) |
|  | Positive for both E and N genes or positive only for N gene with CT 35 or less | 0.004 ( -0.030 , 0.037 ) | 1.02 ( 0.82 , 1.25 ) | 0.59 | * ( -23.2 , 18.6 ) |

\* Doubling/Halving time had an estimated magnitude greater than 50 days and so represented approximately constant prevalence

**Figure 1.**
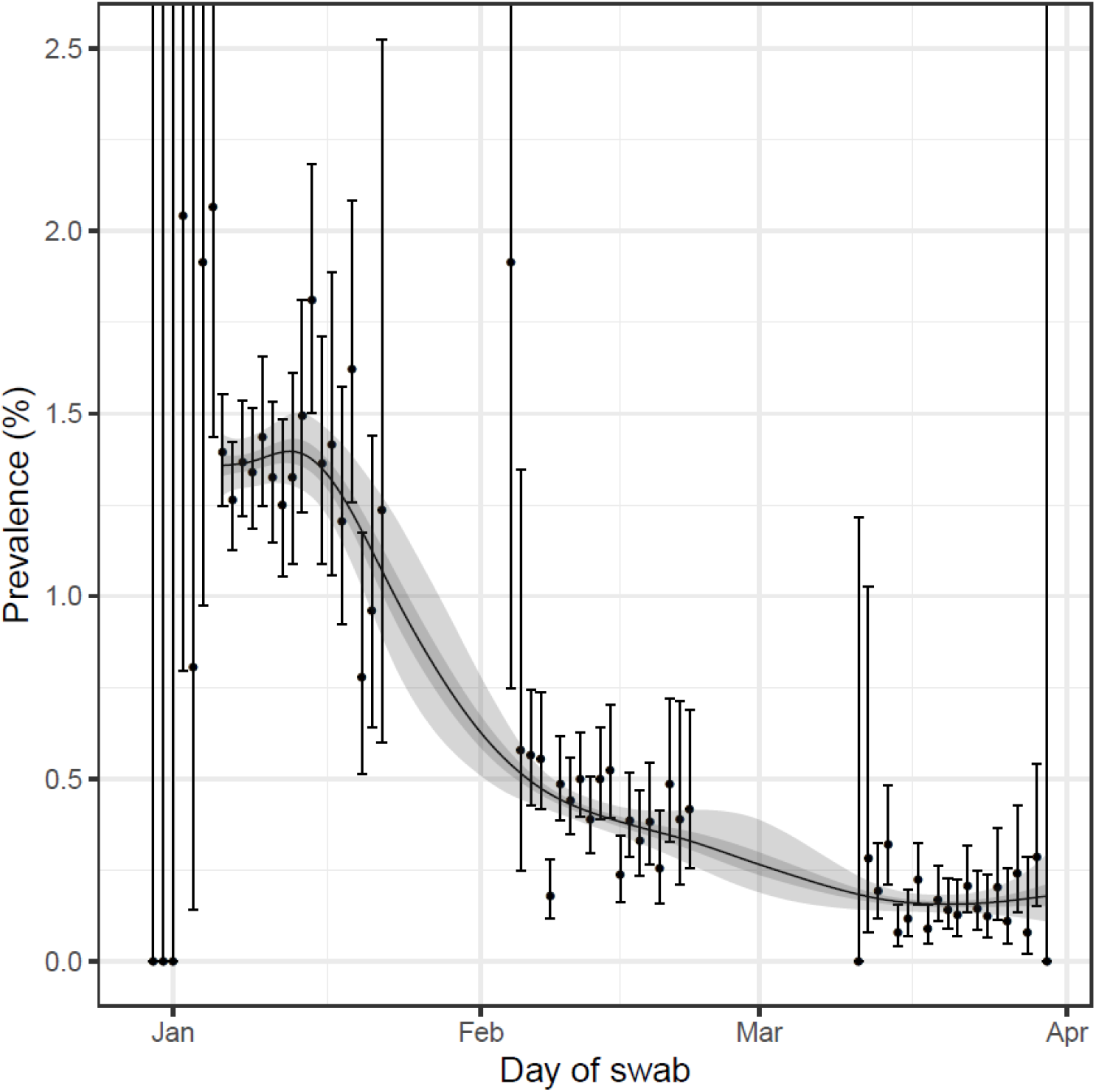
Prevalence of national swab-positivity for England estimated using a P-spline for all nine rounds with central 50% (dark grey) and 95% (light grey) posterior credible intervals. Shown here only for the period of round 8 to round 10. Unweighted observations (black dots) and 95% binomial confidence intervals (vertical lines) are also shown.

At regional level, R over the period of rounds 9 to 10 ranged from 0.75 (0.68, 0.81) in South East to 0.95 (0.87, 1.03) in Yorkshire and The Humber. We estimated a 12% probability that R > 1 between rounds 9 and 10 in Yorkshire and the Humber, while the probability of R >1 was less than 1% in all other regions (Table 3).

**Table 3.** Estimates of regional growth rates, doubling times and reproduction numbers for round 9 to round 10.

| Round | Region | Growth rate | R | Probability R>1 | Halving (-) / Doubling (+) time |
| --- | --- | --- | --- | --- | --- |
| 9 and 10 | East Midlands | -0.026 ( -0.037 , -0.016 ) | 0.84 ( 0.78 , 0.90 ) | <0.01 | -26.4 ( -18.6 , -43.6 ) |
|  | West Midlands | -0.018 ( -0.031 , -0.006 ) | 0.89 ( 0.81 , 0.96 ) | <0.01 | -37.9 ( -22.2 , * ) |
|  | East of England | -0.029 ( -0.042 , -0.018 ) | 0.82 ( 0.75 , 0.89 ) | <0.01 | -23.6 ( -16.5 , -39.2 ) |
|  | London | -0.032 ( -0.044 , -0.020 ) | 0.81 ( 0.74 , 0.88 ) | <0.01 | -22.0 ( -15.7 , -35.2 ) |
|  | North West | -0.026 ( -0.037 , -0.016 ) | 0.84 ( 0.78 , 0.90 ) | <0.01 | -26.6 ( -18.9 , -42.7 ) |
|  | North East | -0.023 ( -0.040 , -0.008 ) | 0.86 ( 0.77 , 0.95 ) | <0.01 | -30.3 ( -17.5 , * ) |
|  | South East | -0.043 ( -0.056 , -0.031 ) | 0.75 ( 0.68 , 0.81 ) | <0.01 | -16.1 ( -12.4 , -22.3 ) |
|  | South West | -0.023 ( -0.041 , -0.007 ) | 0.86 ( 0.76 , 0.96 ) | <0.01 | -30.0 ( -16.9 , * ) |
|  | Yorkshire ** | -0.008 ( -0.022 , 0.005 ) | 0.95 ( 0.87 , 1.03 ) | 0.12 | * ( -32.2 , * ) |

\* Doubling/Halving time had an estimated magnitude greater than 50 days and so represented approximately constant prevalence
\*\* Yorkshire and The Humber

Reflecting these regional R numbers, there were substantial falls in weighted regional prevalence: in South East from 0.36% (0.29%, 0.44%) in round 9 to 0.07% (0.04%, 0.12%) in round 10; London from 0.60% (0.48%, 0.76%) to 0.16% (0.10%, 0.26%); East of England from 0.47% (0.36%, 0.60%) to 0.15% (0.10%, 0.24%); East Midlands from 0.59% (0.45%, 0.77%) to 0.19% (0.13%, 0.28%); and North West from 0.69% (0.54%, 0.88%) to 0.31% (0.21%, 0.45%) (Table 4, Figure 2).

**Table 4a.** Unweighted and weighted prevalence of swab-positivity for sex age and region for rounds 10. *We present weighted prevalence if the number of positives in a category is 10 or more.

| Variable | Category | Round 10 |  |  |  |
| --- | --- | --- | --- | --- | --- |
|  |  | Positive | Total | Unweighted Prevalence | Weighted Prevalence* |
| Gender | Male | 108 | 63,077 | 0.17% ( 0.14% , 0.21% ) | 0.22% ( 0.17% , 0.28% ) |
|  | Female | 119 | 77,765 | 0.15% ( 0.13% , 0.18% ) | 0.18% ( 0.14% , 0.22% ) |
|  | NA | 0 | 2 | 0.00% ( 0.00% , 57.50% ) | NA ( NA , NA ) |
| Age | 05-12 | 33 | 10,750 | 0.31% ( 0.22% , 0.43% ) | 0.41% ( 0.27% , 0.62% ) |
|  | 13-17 | 14 | 7,891 | 0.18% ( 0.11% , 0.30% ) | 0.19% ( 0.11% , 0.34% ) |
|  | 18-24 | 13 | 5,799 | 0.22% ( 0.13% , 0.38% ) | 0.29% ( 0.16% , 0.53% ) |
|  | 25-34 | 21 | 12,203 | 0.17% ( 0.11% , 0.26% ) | 0.18% ( 0.11% , 0.31% ) |
|  | 35-44 | 35 | 17,339 | 0.20% ( 0.15% , 0.28% ) | 0.21% ( 0.14% , 0.31% ) |
|  | 45-54 | 37 | 22,258 | 0.17% ( 0.12% , 0.23% ) | 0.20% ( 0.13% , 0.30% ) |
|  | 55-64 | 39 | 25,811 | 0.15% ( 0.11% , 0.21% ) | 0.17% ( 0.12% , 0.25% ) |
|  | 65-74 | 21 | 25,874 | 0.08% ( 0.05% , 0.12% ) | 0.09% ( 0.05% , 0.16% ) |
|  | 75+ | 14 | 12,919 | 0.11% ( 0.06% , 0.18% ) | 0.09% ( 0.05% , 0.16% ) |
| Region | South East | 18 | 31,128 | 0.06% ( 0.04% , 0.09% ) | 0.07% ( 0.04% , 0.12% ) |
|  | North East | 16 | 4,935 | 0.32% ( 0.20% , 0.53% ) | 0.41% ( 0.24% , 0.70% ) |
|  | North West | 41 | 16,106 | 0.25% ( 0.19% , 0.35% ) | 0.31% ( 0.21% , 0.45% ) |
|  | Yorkshire and The Humber | 28 | 9,601 | 0.29% ( 0.20% , 0.42% ) | 0.36% ( 0.22% , 0.57% ) |
|  | East Midlands | 34 | 17,934 | 0.19% ( 0.14% , 0.26% ) | 0.19% ( 0.13% , 0.28% ) |
|  | West Midlands | 26 | 12,926 | 0.20% ( 0.14% , 0.29% ) | 0.20% ( 0.13% , 0.31% ) |
|  | East of England | 27 | 20,103 | 0.13% ( 0.09% , 0.20% ) | 0.15% ( 0.10% , 0.24% ) |
|  | London | 22 | 14,062 | 0.16% ( 0.10% , 0.24% ) | 0.16% ( 0.10% , 0.26% ) |
|  | South West | 15 | 14,049 | 0.11% ( 0.06% , 0.18% ) | 0.13% ( 0.07% , 0.26% ) |
| Employment type | Health care or care home worker | 19 | 8,614 | 0.22% ( 0.14% , 0.34% ) | 0.21% ( 0.12% , 0.36% ) |
|  | Other essential/key worker | 44 | 24,094 | 0.18% ( 0.14% , 0.25% ) | 0.20% ( 0.14% , 0.30% ) |
|  | Other worker | 90 | 52,495 | 0.17% ( 0.14% , 0.21% ) | 0.23% ( 0.18% , 0.30% ) |
|  | Not full-time, part-time, or self-employed | 64 | 52,331 | 0.12% ( 0.10% , 0.16% ) | 0.14% ( 0.11% , 0.19% ) |
|  | NA | 10 | 3,310 | 0.30% ( 0.16% , 0.56% ) | 0.33% ( 0.17% , 0.64% ) |
| Ethnic group | White | 194 | 126,966 | 0.15% ( 0.13% , 0.18% ) | 0.19% ( 0.16% , 0.23% ) |
|  | Asian | 14 | 5,892 | 0.24% ( 0.14% , 0.40% ) | 0.27% ( 0.15% , 0.47% ) |
|  | Black | 4 | 1,965 | 0.20% ( 0.08% , 0.52% ) | 0.11% ( 0.04% , 0.31% ) |
|  | Mixed | 7 | 2,460 | 0.28% ( 0.14% , 0.59% ) | 0.32% ( 0.15% , 0.71% ) |
|  | Other | 3 | 1,143 | 0.26% ( 0.09% , 0.77% ) | 0.15% ( 0.04% , 0.47% ) |
|  | NA | 5 | 2,418 | 0.21% ( 0.09% , 0.48% ) | 0.22% ( 0.07% , 0.62% ) |
| Household size | 1 | 26 | 20,336 | 0.13% ( 0.09% , 0.19% ) | 0.15% ( 0.09% , 0.24% ) |
|  | 2 | 64 | 53,128 | 0.12% ( 0.09% , 0.15% ) | 0.16% ( 0.11% , 0.22% ) |
|  | 3 | 48 | 25,373 | 0.19% ( 0.14% , 0.25% ) | 0.22% ( 0.15% , 0.33% ) |
|  | 4 | 58 | 28,568 | 0.20% ( 0.16% , 0.26% ) | 0.24% ( 0.18% , 0.33% ) |
|  | 5 | 19 | 9,582 | 0.20% ( 0.13% , 0.31% ) | 0.21% ( 0.11% , 0.38% ) |
|  | 6+ | 12 | 3,857 | 0.31% ( 0.18% , 0.54% ) | 0.35% ( 0.19% , 0.65% ) |
| COVID case contact | No | 136 | 119,963 | 0.11% ( 0.10% , 0.13% ) | 0.14% ( 0.11% , 0.17% ) |
|  | Yes, contact with a confirmed/tested COVID-19 case | 46 | 977 | 4.71% ( 3.55% , 6.22% ) | 4.86% ( 3.37% , 6.96% ) |
|  | Yes, contact with a suspected COVID-19 case | 5 | 397 | 1.26% ( 0.54% , 2.91% ) | 0.90% ( 0.32% , 2.48% ) |
|  | NA | 40 | 19,507 | 0.21% ( 0.15% , 0.28% ) | 0.27% ( 0.18% , 0.40% ) |
| Symptom status | Classic COVID symptoms | 45 | 3,508 | 1.28% ( 0.96% , 1.71% ) | 1.56% ( 1.09% , 2.24% ) |
|  | Other symptoms | 30 | 14,801 | 0.20% ( 0.14% , 0.29% ) | 0.28% ( 0.18% , 0.42% ) |
|  | No symptoms | 112 | 103,135 | 0.11% ( 0.09% , 0.13% ) | 0.12% ( 0.10% , 0.15% ) |
|  | NA | 40 | 19,400 | 0.21% ( 0.15% , 0.28% ) | 0.27% ( 0.18% , 0.40% ) |
| Deprivation | 1 Most deprived | 36 | 12,974 | 0.28% ( 0.20% , 0.38% ) | 0.33% ( 0.23% , 0.47% ) |
|  | 2 | 46 | 21,750 | 0.21% ( 0.16% , 0.28% ) | 0.25% ( 0.18% , 0.35% ) |
|  | 3 | 44 | 30,461 | 0.14% ( 0.11% , 0.19% ) | 0.17% ( 0.12% , 0.24% ) |
|  | 4 | 48 | 35,151 | 0.14% ( 0.10% , 0.18% ) | 0.13% ( 0.09% , 0.17% ) |
|  | 5 Least deprived | 53 | 40,508 | 0.13% ( 0.10% , 0.17% ) | 0.13% ( 0.09% , 0.17% ) |

**Table 4b.**
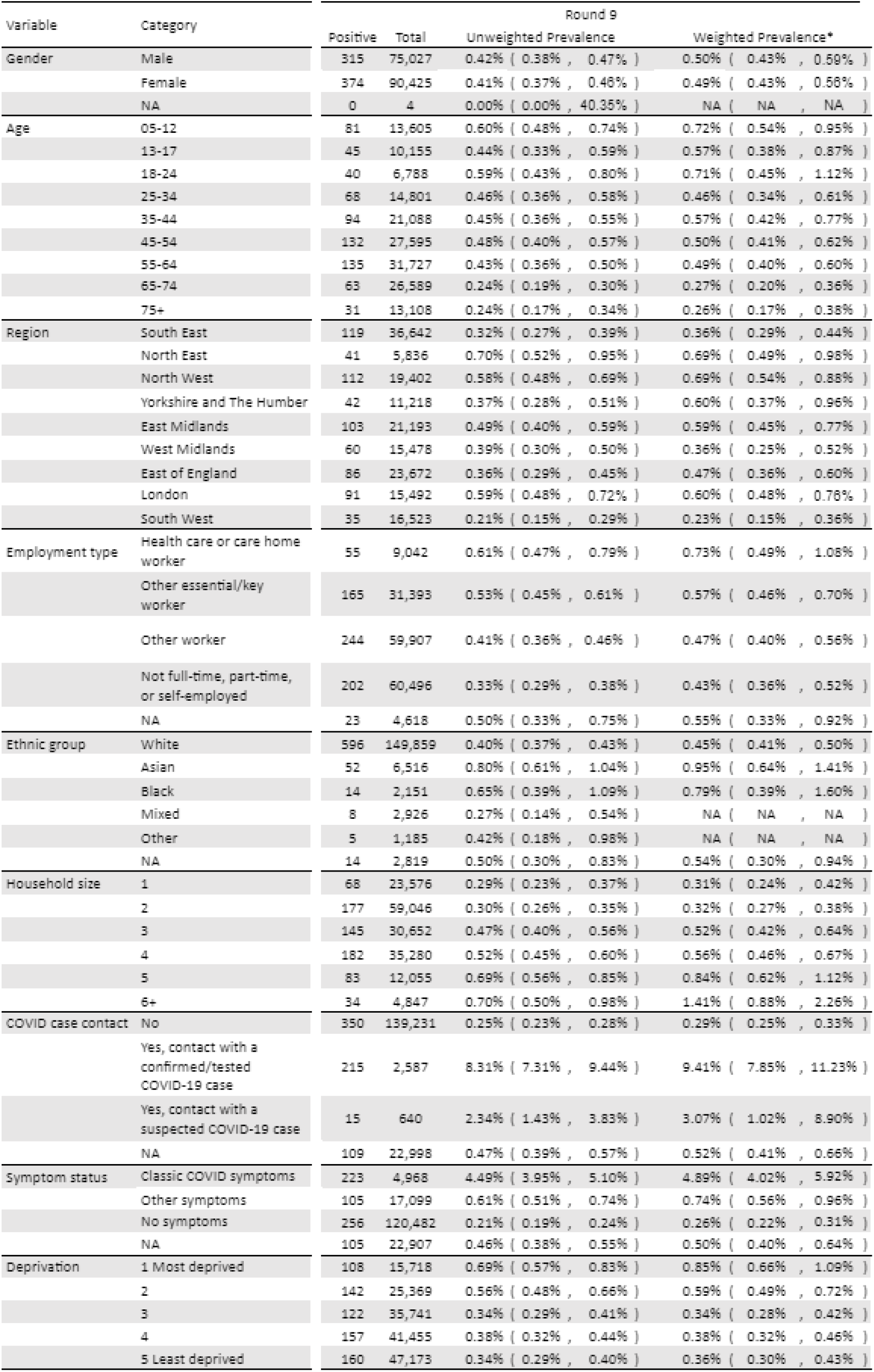
Unweighted and weighted prevalence of swab-positivity for sex age and region for rounds 9. *We present weighted prevalence if the number of positives in a category is 10 or more.

| Variable | Category | Round 9 |  |  |  |
| --- | --- | --- | --- | --- | --- |
|  |  | Positive | Total | Unweighted Prevalence | Weighted Prevalence* |
| Gender | Male | 315 | 75,027 | 0.42% ( 0.38% , 0.47% ) | 0.50% ( 0.43% , 0.59% ) |
|  | Female | 374 | 90,425 | 0.41% ( 0.37% , 0.46% ) | 0.49% ( 0.43% , 0.56% ) |
|  | NA | 0 | 4 | 0.00% ( 0.00% , 40.35% ) | NA ( NA , NA ) |
| Age | 05-12 | 81 | 13,605 | 0.60% ( 0.48% , 0.74% ) | 0.72% ( 0.54% , 0.95% ) |
|  | 13-17 | 45 | 10,155 | 0.44% ( 0.33% , 0.59% ) | 0.57% ( 0.38% , 0.87% ) |
|  | 18-24 | 40 | 6,788 | 0.59% ( 0.43% , 0.80% ) | 0.71% ( 0.45% , 1.12% ) |
|  | 25-34 | 68 | 14,801 | 0.46% ( 0.36% , 0.58% ) | 0.46% ( 0.34% , 0.61% ) |
|  | 35-44 | 94 | 21,088 | 0.45% ( 0.36% , 0.55% ) | 0.57% ( 0.42% , 0.77% ) |
|  | 45-54 | 132 | 27,595 | 0.48% ( 0.40% , 0.57% ) | 0.50% ( 0.41% , 0.62% ) |
|  | 55-64 | 135 | 31,727 | 0.43% ( 0.36% , 0.50% ) | 0.49% ( 0.40% , 0.60% ) |
|  | 65-74 | 63 | 26,589 | 0.24% ( 0.19% , 0.30% ) | 0.27% ( 0.20% , 0.36% ) |
|  | 75+ | 31 | 13,108 | 0.24% ( 0.17% , 0.34% ) | 0.26% ( 0.17% , 0.38% ) |
| Region | South East | 119 | 36,642 | 0.32% ( 0.27% , 0.39% ) | 0.36% ( 0.29% , 0.44% ) |
|  | North East | 41 | 5,836 | 0.70% ( 0.52% , 0.95% ) | 0.69% ( 0.49% , 0.98% ) |
|  | North West | 112 | 19,402 | 0.58% ( 0.48% , 0.69% ) | 0.69% ( 0.54% , 0.88% ) |
|  | Yorkshire and The Humber | 42 | 11,218 | 0.37% ( 0.28% , 0.51% ) | 0.60% ( 0.37% , 0.96% ) |
|  | East Midlands | 103 | 21,193 | 0.49% ( 0.40% , 0.59% ) | 0.59% ( 0.45% , 0.77% ) |
|  | West Midlands | 60 | 15,478 | 0.39% ( 0.30% , 0.50% ) | 0.36% ( 0.25% , 0.52% ) |
|  | East of England | 86 | 23,672 | 0.36% ( 0.29% , 0.45% ) | 0.47% ( 0.36% , 0.60% ) |
|  | London | 91 | 15,492 | 0.59% ( 0.48% , 0.72% ) | 0.60% ( 0.48% , 0.76% ) |
|  | South West | 35 | 16,523 | 0.21% ( 0.15% , 0.29% ) | 0.23% ( 0.15% , 0.36% ) |
| Employment type | Health care or care home worker | 55 | 9,042 | 0.61% ( 0.47% , 0.79% ) | 0.73% ( 0.49% , 1.08% ) |
|  | Other essential/key worker | 165 | 31,393 | 0.53% ( 0.45% , 0.61% ) | 0.57% ( 0.46% , 0.70% ) |
|  | Other worker | 244 | 59,907 | 0.41% ( 0.36% , 0.46% ) | 0.47% ( 0.40% , 0.56% ) |
|  | Not full-time, part-time, or self-employed | 202 | 60,496 | 0.33% ( 0.29% , 0.38% ) | 0.43% ( 0.36% , 0.52% ) |
|  | NA | 23 | 4,618 | 0.50% ( 0.33% , 0.75% ) | 0.55% ( 0.33% , 0.92% ) |
| Ethnic group | White | 596 | 149,859 | 0.40% ( 0.37% , 0.43% ) | 0.45% ( 0.41% , 0.50% ) |
|  | Asian | 52 | 6,516 | 0.80% ( 0.61% , 1.04% ) | 0.95% ( 0.64% , 1.41% ) |
|  | Black | 14 | 2,151 | 0.65% ( 0.39% , 1.09% ) | 0.79% ( 0.39% , 1.60% ) |
|  | Mixed | 8 | 2,926 | 0.27% ( 0.14% , 0.54% ) | NA ( NA , NA ) |
|  | Other | 5 | 1,185 | 0.42% ( 0.18% , 0.98% ) | NA ( NA , NA ) |
|  | NA | 14 | 2,819 | 0.50% ( 0.30% , 0.83% ) | 0.54% ( 0.30% , 0.94% ) |
| Household size | 1 | 68 | 23,576 | 0.29% ( 0.23% , 0.37% ) | 0.31% ( 0.24% , 0.42% ) |
|  | 2 | 177 | 59,046 | 0.30% ( 0.26% , 0.35% ) | 0.32% ( 0.27% , 0.38% ) |
|  | 3 | 145 | 30,652 | 0.47% ( 0.40% , 0.56% ) | 0.52% ( 0.42% , 0.64% ) |
|  | 4 | 182 | 35,280 | 0.52% ( 0.45% , 0.60% ) | 0.56% ( 0.46% , 0.67% ) |
|  | 5 | 83 | 12,055 | 0.69% ( 0.56% , 0.85% ) | 0.84% ( 0.62% , 1.12% ) |
|  | 6+ | 34 | 4,847 | 0.70% ( 0.50% , 0.98% ) | 1.41% ( 0.88% , 2.26% ) |
| COVID case contact | No | 350 | 139,231 | 0.25% ( 0.23% , 0.28% ) | 0.29% ( 0.25% , 0.33% ) |
|  | Yes, contact with a confirmed/tested COVID-19 case | 215 | 2,587 | 8.31% ( 7.31% , 9.44% ) | 9.41% ( 7.85% , 11.23% ) |
|  | Yes, contact with a suspected COVID-19 case | 15 | 640 | 2.34% ( 1.43% , 3.83% ) | 3.07% ( 1.02% , 8.90% ) |
|  | NA | 109 | 22,998 | 0.47% ( 0.39% , 0.57% ) | 0.52% ( 0.41% , 0.66% ) |
| Symptom status | Classic COVID symptoms | 223 | 4,968 | 4.49% ( 3.95% , 5.10% ) | 4.89% ( 4.02% , 5.92% ) |
|  | Other symptoms | 105 | 17,099 | 0.61% ( 0.51% , 0.74% ) | 0.74% ( 0.56% , 0.96% ) |
|  | No symptoms | 256 | 120,482 | 0.21% ( 0.19% , 0.24% ) | 0.26% ( 0.22% , 0.31% ) |
|  | NA | 105 | 22,907 | 0.46% ( 0.38% , 0.55% ) | 0.50% ( 0.40% , 0.64% ) |
| Deprivation | 1 Most deprived | 108 | 15,718 | 0.69% ( 0.57% , 0.83% ) | 0.85% ( 0.66% , 1.09% ) |
|  | 2 | 142 | 25,369 | 0.56% ( 0.48% , 0.66% ) | 0.59% ( 0.49% , 0.72% ) |
|  | 3 | 122 | 35,741 | 0.34% ( 0.29% , 0.41% ) | 0.34% ( 0.28% , 0.42% ) |
|  | 4 | 157 | 41,455 | 0.38% ( 0.32% , 0.44% ) | 0.38% ( 0.32% , 0.46% ) |
|  | 5 Least deprived | 160 | 47,173 | 0.34% ( 0.29% , 0.40% ) | 0.36% ( 0.30% , 0.43% ) |

**Figure 2.**
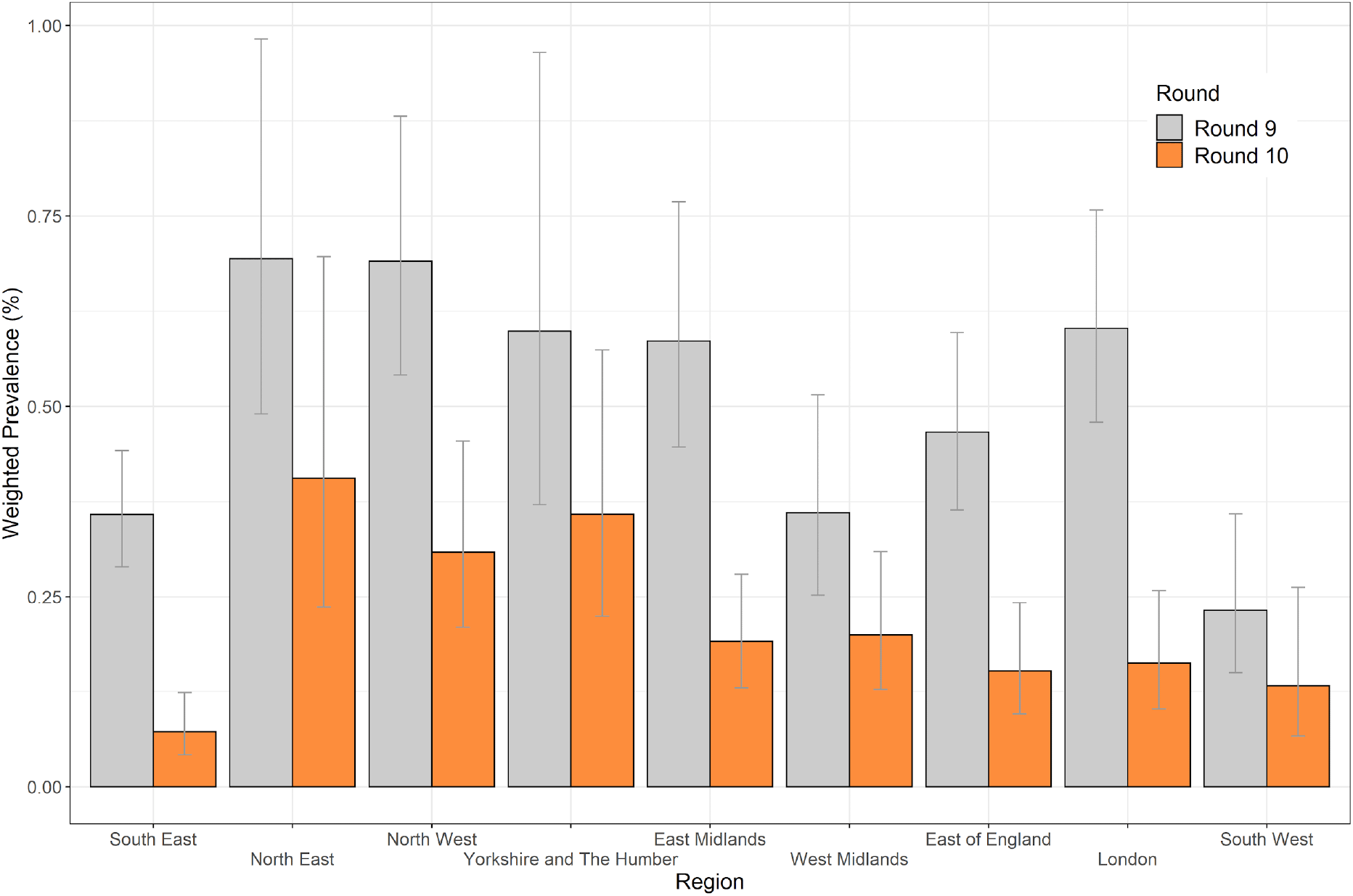
Weighted prevalence of swab-positivity by region for rounds 9 and 10. Bars show 95% confidence intervals. See Table 4.

At sub-regional scale, areas of apparent higher prevalence remain in round 10 in parts of North West, and Yorkshire and The Humber (Figure 3).

**Figure 3.**
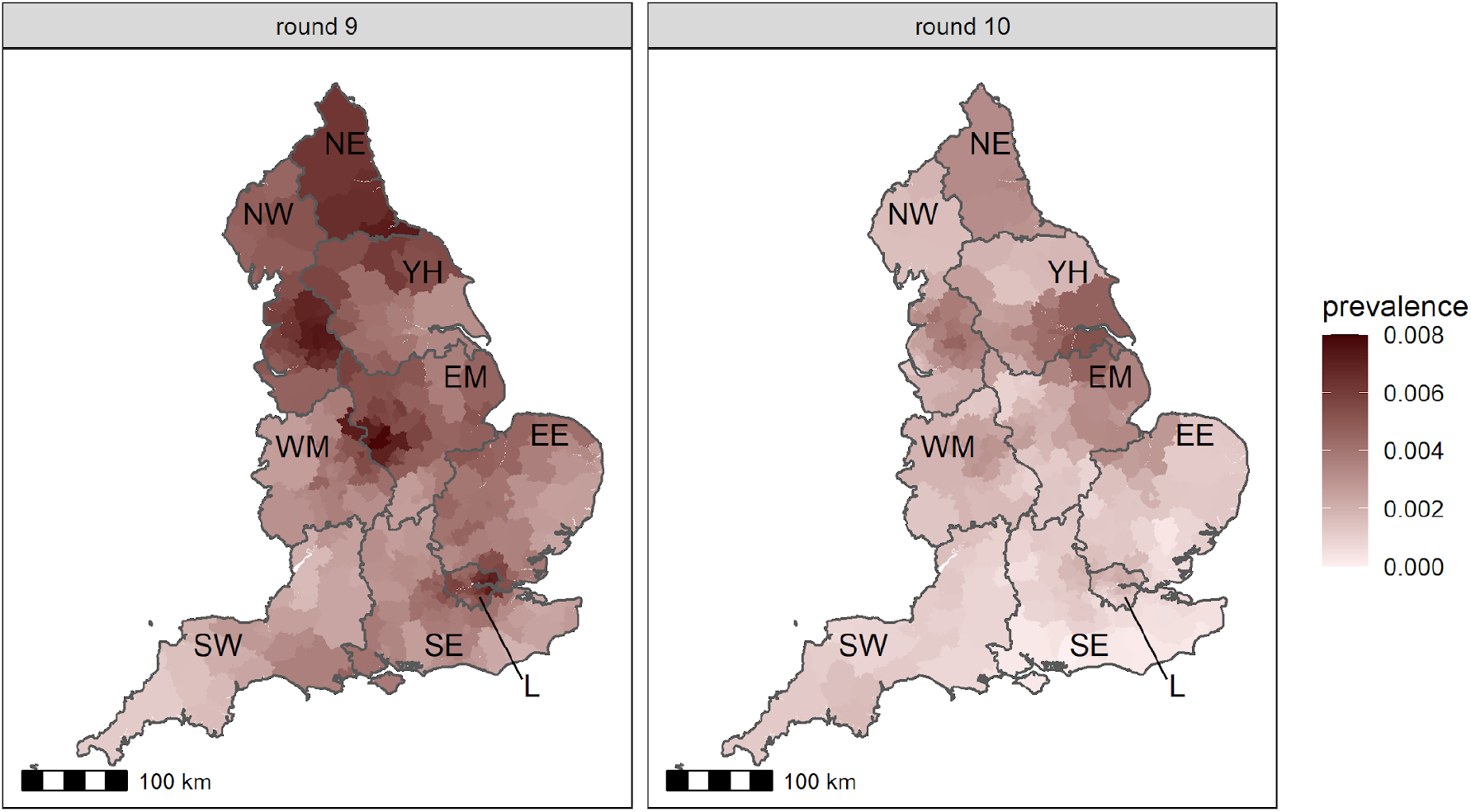
Neighbourhood prevalence of swab-positivity for rounds 9 and 10. Neighbourhood prevalence calculated from 4,732 (round 9) and 4,016 (round 10) nearest neighbours in the sample (the median number of neighbours within 30km in the study). Neighbourhood prevalence displayed as an average of a random sample of 15 participants for each lower-tier local authority (LTLA). Regions: NE = North East, NW = North West, YH = Yorkshire and The Humber, EM = East Midlands, WM = West Midlands, EE = East of England, L = London, SE = South East, SW = South West.

We found the highest weighted prevalence of swab-positivity in those aged 5 to 12 years at 0.41% (0.27%, 0.62%) compared with the lowest in those aged 65 to 74 years and 75 years and over at 0.09% (0.05%, 0.16%) (Table 4, Figure 4). As with prior rounds, we found much higher prevalence of swab-positivity during round 10 in those who had come into contact with a confirmed COVID-19 case at 4.9% (3.4%, 7.0%) compared with those who had not at 0.14% (0.11%, 0.17%) (Table 4a). Also, those living in the least deprived neighbourhoods had a lower prevalence of swab-positivity at 0.13% (0.09%, 0.17%) compared with 0.33% (0.23%, 0.47%) for those in the most deprived neighbourhoods (Table 4a).

**Figure 4.**
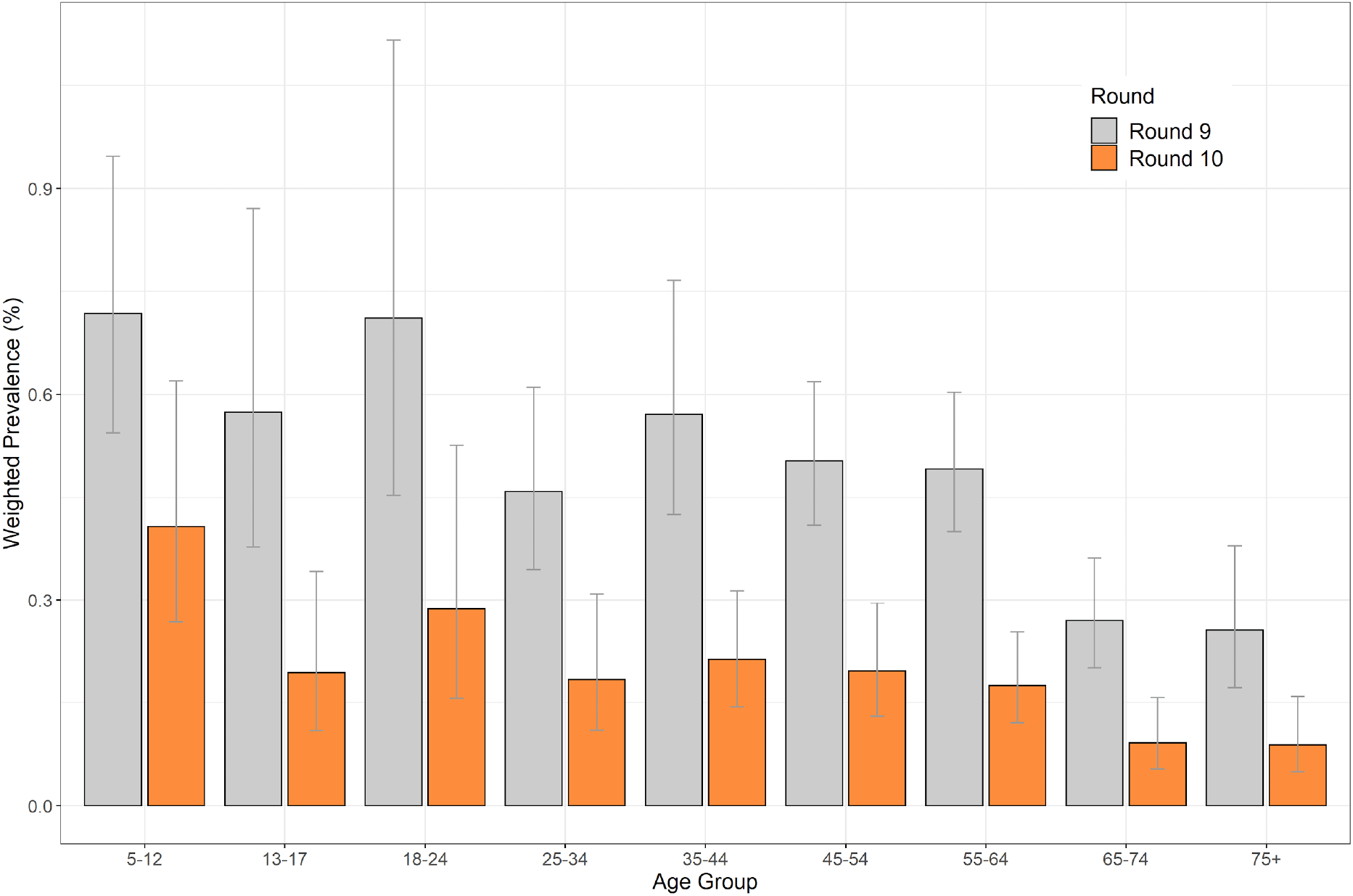
Weighted prevalence of swab-positivity by age groups for rounds 9 and 10. Bars show 95% confidence intervals. See Table 4.

Frequency of foreign travel remained low overall during round 10 compared with prior rounds of REACT-1 (Figure 5). Foreign travel remains higher in London compared with other regions except for Yorkshire and The Humber.

**Figure 5.**
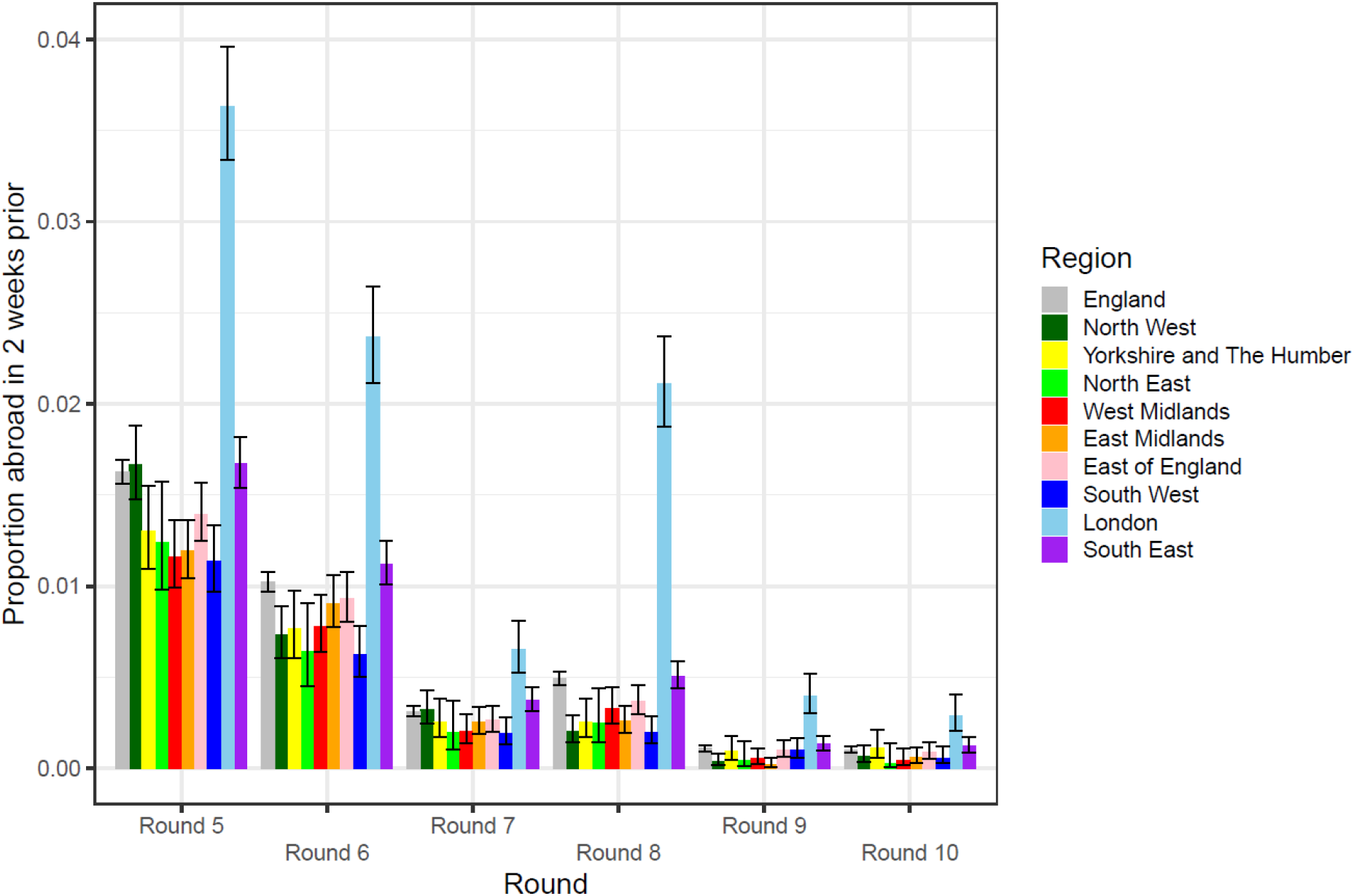
Unweighted proportion of people who had travelled abroad in the previous two weeks in England and its nine constituent regions for rounds 5 to 10. Vertical line shows 95% binomial confidence intervals.

We investigated the relationship between swab-positivity, as estimated in rounds 1 to 8 of REACT-1, and hospital admissions (Figure 6), and deaths (Figure 7) from publicly available national data. We estimated lags of 18 (18, 18) days between the timing of swab results and hospital admissions, and 27 (27, 27) days between swab and death to give the best fit to REACT-1 rounds 1 to 8 prevalence data. During the period of round 9, curves for deaths and (more marginally) for hospital admissions diverged from the prevalence P-spline, suggesting that infections may have resulted in fewer hospitalisations and deaths since the start of widespread vaccination.

**Figure 6.**
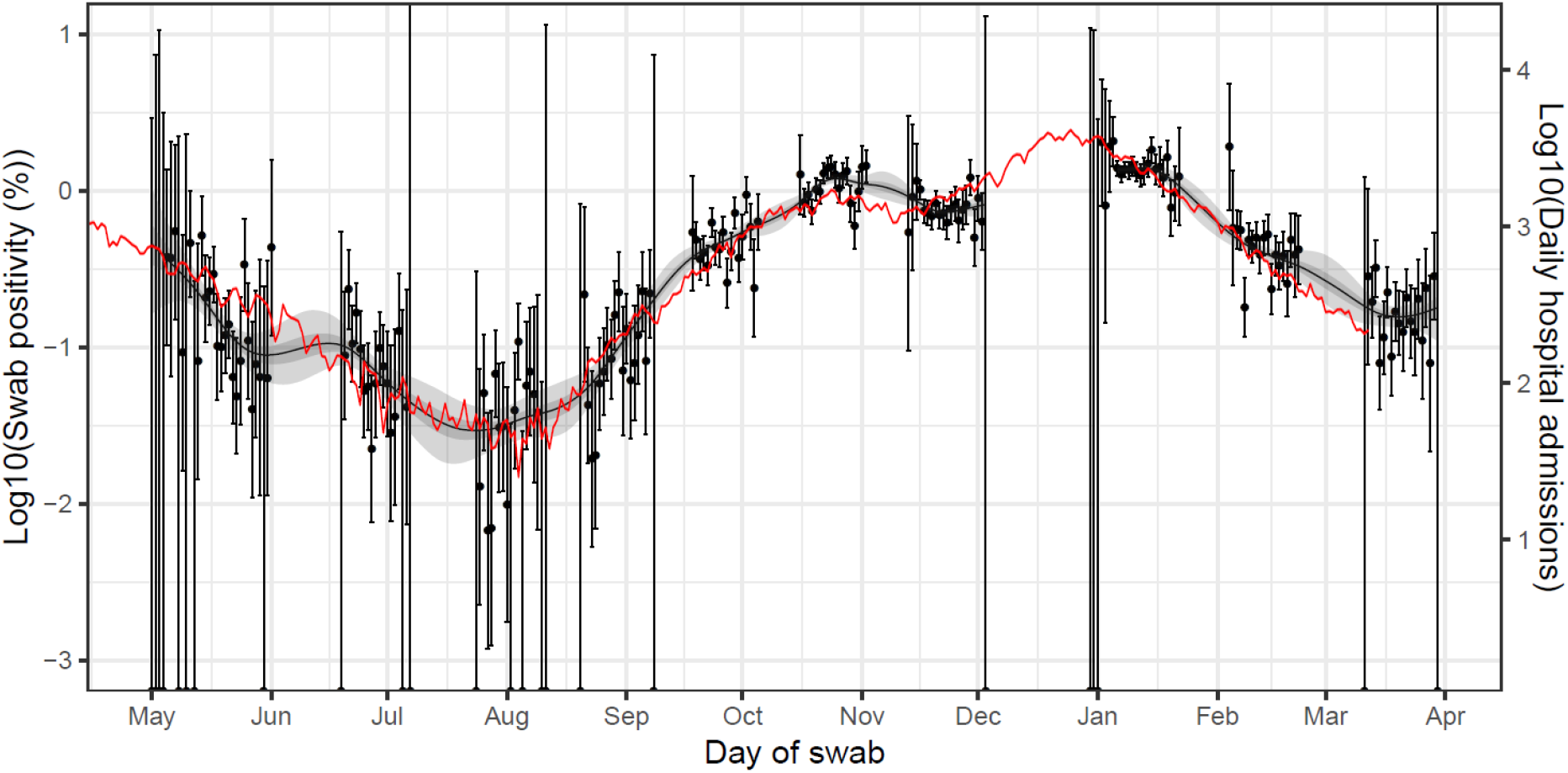
Daily hospital admissions in England (solid red line, right hand y-axis) shifted by a lag parameter along the x-axis (see below), and daily swab-positivity for all 9 rounds of the study (dots with 95% confidence intervals, left hand y-axis) and the P-spline estimate for swab positivity (Solid black line, dark grey shaded area is 50% central credible interval, light grey shaded area is 95% central credible interval, left-hand y-axis). Daily hospital admissions have been fit to observations from the REACT-1 rounds 1-8 to obtain scaling and lag parameters. These parameter values were estimated using a Bayesian MCMC model: daily_positives(t) ∼ Binomial(daily_swab_tests(t), p = daily_admissions(t+lag)*scale). The discrete time lag parameter was estimated at 18 (18, 18) days. Note the P-spline is not plotted for the region between round 7 and 8 in which there was a peak in swab-positivity while REACT-1 was not active.

**Figure 7.**
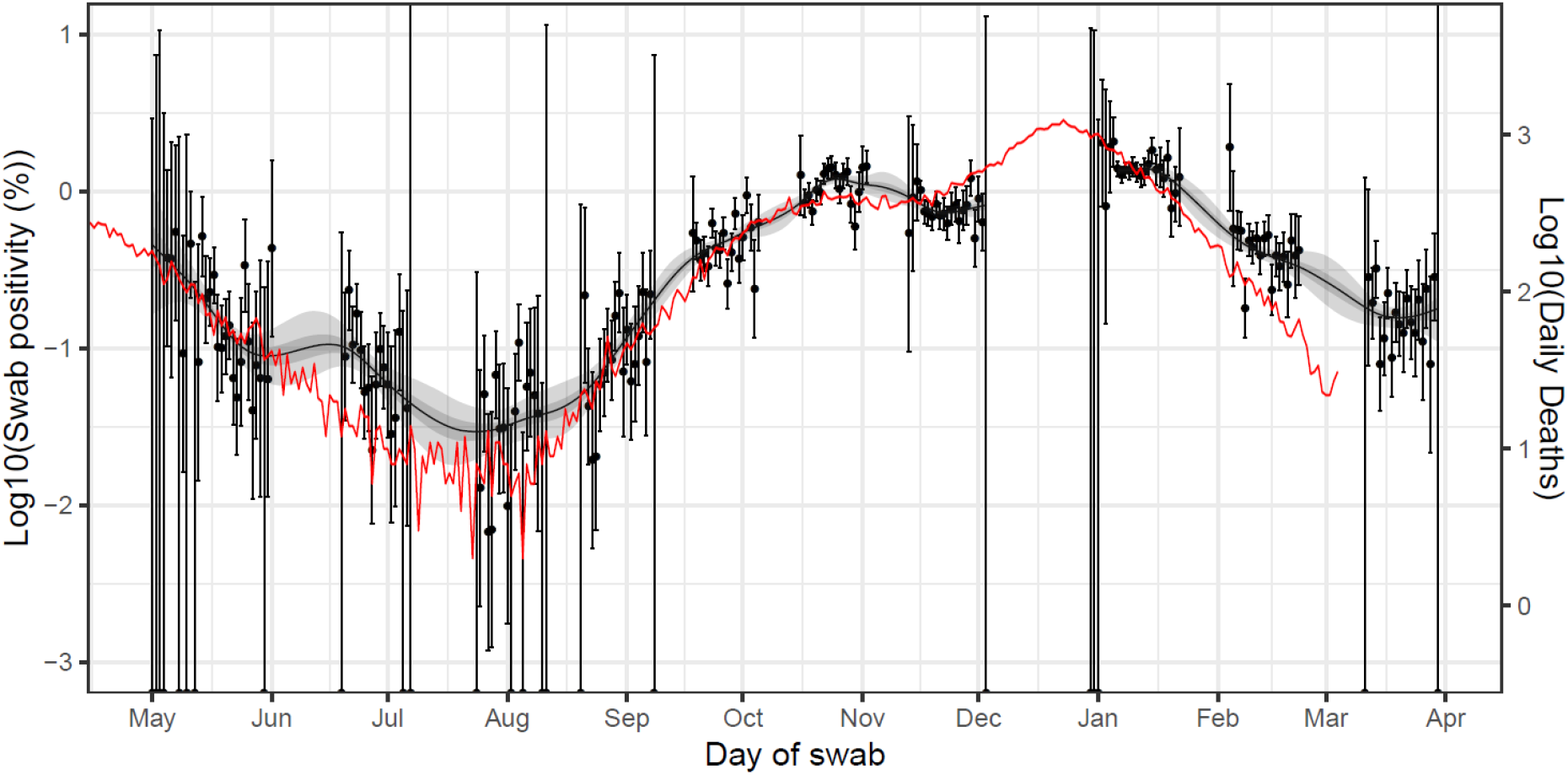
Daily deaths in England (solid red line, right hand y-axis) shifted by a lag parameter along the x-axis (see below), and daily swab positivity for all 9 rounds of the study (dots with 95% confidence intervals, left hand y-axis) and the P-spline estimate for swab positivity (Solid black line, dark grey shaded area is 50% central credible interval, light grey shaded area is 95% central credible interval, left-hand y-axis). Daily deaths have been fit to observations from the REACT-1 rounds 1-8 to obtain scaling and lag parameters. These parameter values were estimated using a Bayesian MCMC model: daily_positives(t) ∼ Binomial(daily_swab_tests(t), p = daily_admissions(t+lag)*scale). The discrete time lag parameter was estimated at 27 (27, 27) days. Note the P-spline is not plotted for the region between round 7 and 8 in which there was a peak in swab-positivity while REACT-1 was not active.

## Discussion

In this tenth round of the REACT-1 study carried out from 11 to 30 March 2021, we report an approximate 60% reduction in prevalence since the previous round from 4 to 23 February. These rounds occurred during the third national lockdown in England [6] following the second wave of infections, which grew rapidly through December 2020 until the beginning of January 2021. The most recent round also covered the start of the gradual easing of lockdown [3], notably the opening up of schools from 8 March 2021.

Our data indicate a slowing of the epidemic from February to March, with the national R number over that period reliably below one. However, in the most recent data for March, the rate of decline slowed and prevalence flattened at 1 in 500 (0.20%) people infected overall, with prevalence highest in primary and early secondary school-aged children (5 to 12 years) and lowest in those above the age of 65 years. The lower rate among older adults is consistent with an effect from the vaccination roll-out in England, that has focused initially on older people and the most vulnerable.

Our analysis of mortality and to a lesser extent hospitalisation trends is consistent with the vaccination programme contributing in part to lower rates of severe outcomes in the older age group. In recent rounds of REACT-1 there was a close approximation between prevalence of infection (measured by REACT-1) and hospitalisation and mortality rates (from publicly available national data sources, after considering a suitable lag period between infection and outcome). This relationship has diverged, particularly for deaths, from late January onwards and is in keeping with the timing of the roll-out of the mass vaccination programme. This observation is consistent with studies in Scotland [7] and Israel [8] which reported high efficacy of vaccination against severe disease. It will be important to monitor future trends in the relationship between infections, deaths and hospitalisations in England. Specifically if the approximation of the infection and mortality curves revert to prior patterns, this might indicate an increase in frequency of variants associated with reduced efficacy of the vaccine. We note that during early rounds of REACT-1 the relationship between infections and mortality appeared weaker: this likely reflects a higher proportion of infections occurring in health and care facilities at that time rather than in the community later in the epidemic [9].

Swab-positivity was over 30-fold higher in participants who reported contact with a confirmed case compared with those who did not. This reinforces the high potential value of testing known contacts even though manual contact tracing alone may not be sufficient to achieve R < 1 in the absence of other interventions [10,11]. Further, the consistently higher rates of infection in the most deprived neighbourhoods observed in REACT-1 suggest a potential role for targeted highly-local interventions during periods of lower prevalence, even when the presence of a novel variant is not a primary concern [12].

Prevalence of infection in England is now at a level last seen in early September 2020, and nearing the prevalence at the end of the first lockdown in England in May 2020. The Government in England has adopted a cautionary approach to easing of lockdown with approximately five week intervals between each step toward ‘normalisation’ of activities including opening of non-essential shops, indoor sports, outdoor hospitality and personal services (such as hairdressing) from 12 April 2021 [3]. This approach allows time to assess the effect of each step on the transmission of the virus and the R number, which will be measured in subsequent REACT-1 surveys, among others, over the ensuing period. Meanwhile, our estimate of 1.0 for the R number in March suggests that the fine balance between the cautious opening up of society and the need to control the levels of infection has not so far resulted in a third wave of infections. The continued roll-out of the vaccination programme, including second doses, and the extension to adult ages below 50 years will further increase immunity in the population.

## Methods

REACT-1 involves a random sample of the population in England (ages 5 years and over) who are invited to undertake a self-administered throat and nose swab in the home (parent/guardian administered at ages 5 to 12 years) and complete a health and lifestyle questionnaire [5]. We request that completed swabs are refrigerated before being transported by courier and sent chilled to the laboratory for RT-PCR testing.

We obtain the sample using the National Health Service patient register and aim to achieve approximately equal numbers of participants in each of the 315 lower tier local authorities (LTLAs) in England. In round 10, we sent out 760,000 letters to named individuals and dispatched 180,106 (23.7%) test kits, resulting in 140,844 (78.2%) completed swabs with a valid test result. This gave an overall response rate (valid swabs divided by total number of letters sent) of 18.5%.

We obtain prevalence of RT-PCR swab-positivity with and without weighting. This is done both to take account of the sample design (balanced by LTLA, not by population) and to adjust for variable non-response. The aim is to provide prevalence estimates (national, regional, and by specific demographic and occupational groups) that are representative of the population of England as a whole, by age, sex, region and ethnicity. We also obtain a map of smoothed prevalence at LTLA level by use of a neighbourhood spatial smoothing method across nearest neighbours up to 30 km.

We use exponential growth models to estimate the reproduction number R, across and within rounds. Across all rounds, we fit a smoothed P-spline function to the daily prevalence data, with knots at 5-day intervals, to examine trends in unweighted prevalence over time [13]. We compare the daily prevalence data in REACT-1 with the publicly available national daily hospital admissions and COVID-19 mortality data (deaths within 28 days of a positive test), with allowance for a suitable discrete-day lag period between infection and either hospital admission or death and an appropriate scaling parameter.

In sensitivity analyses for the estimation of R, we provide alternative cut-points of cycle threshold (CT) values for swab-positivity and restrict the analyses only to those who did not report symptoms in the previous week.

Statistical analyses were carried out in R [14]. We obtained research ethics approval from the South Central-Berkshire B Research Ethics Committee (IRAS ID: 283787).

## Data Availability

Links to a spreadsheet, and the GitHub R pacakge containing supporting data for tables and figures are available within the manuscript

## Data availability

Supporting data for tables and figures are available either: in this [spreadsheet](https://docs.google.com/spreadsheets/d/1EPiv49Twk9z4vtH3iYAJEn1swRZL_oobIxpn3nSlauM/edit#gid=1192958503); or in the inst/extdata directory of this [GitHub R package](https://github.com/mrc-ide/reactidd).

## Declaration of interests

We declare no competing interests.

## Funding

The study was funded by the Department of Health and Social Care in England.

## Acknowledgements

SR, CAD acknowledge support: MRC Centre for Global Infectious Disease Analysis, National Institute for Health Research (NIHR) Health Protection Research Unit (HPRU), Wellcome Trust (200861/Z/16/Z, 200187/Z/15/Z), and Centres for Disease Control and Prevention (US, U01CK0005-01-02). GC is supported by an NIHR Professorship. HW acknowledges support from an NIHR Senior Investigator Award and the Wellcome Trust (205456/Z/16/Z). PE is Director of the MRC Centre for Environment and Health (MR/L01341X/1, MR/S019669/1). PE acknowledges support from Health Data Research UK (HDR UK); the NIHR Imperial Biomedical Research Centre; NIHR HPRUs in Chemical and Radiation Threats and Hazards, and Environmental Exposures and Health; the British Heart Foundation Centre for Research Excellence at Imperial College London (RE/18/4/34215); and the UK Dementia Research Institute at Imperial (MC_PC_17114). We thank The Huo Family Foundation for their support of our work on COVID-19.

We thank key collaborators on this work – Ipsos MORI: Kelly Beaver, Sam Clemens, Gary Welch, Nicholas Gilby, Kelly Ward and Kevin Pickering; Institute of Global Health Innovation at Imperial College: Gianluca Fontana, Sutha Satkunarajah, Didi Thompson and Lenny Naar; Molecular Diagnostic Unit, Imperial College London: Prof. Graham Taylor; North West London Pathology and Public Health England for help in calibration of the laboratory analyses; Patient Experience Research Centre at Imperial College and the REACT Public Advisory Panel; NHS Digital for access to the NHS register; and the Department of Health and Social Care for logistic support. SR acknowledges helpful discussion with attendees of meetings of the UK Government Office for Science (GO-Science) Scientific Pandemic Influenza – Modelling (SPI-M) committee.

## Notes

### Competing Interest Statement

The authors have declared no competing interest.

### Author Declarations

We obtained research ethics approval from the South Central-Berkshire B Research Ethics Committee (IRAS ID: 283787).

